# Developing and validating a knowledge and skills test for Tanzanian birth attendants trained in PartoMa safe childbirth care

**DOI:** 10.1101/2025.02.26.25322943

**Authors:** Rashid Saleh Khamis, D. van Herwaarden, Natasha Housseine, Tarek Meguid, Nanna Maaløe, Jos van Roosmalen, Monica Lauridsen Kujabi, Brenda Sequeira Dmello, Mikael Henriksen, Suhaila Salum Yussuf, Salma Abdi Mahmoud, Karl Bang Christensen, Ib Christian Bygbjerg, Thomas van den Akker, Dan Wolf Meyrowitsch

**Author notes:** Corresponding Author**.(RSK)**.

## Abstract

**Introduction:** Improving quality of maternity care in low- and middle-income countries is essential for reducing maternal and perinatal mortality and morbidity. Low dose-high frequency in-house training in routine and emergency maternity care is suggested to be central for this. To ensure the effectiveness and resource-efficiency of such training, a knowledge and skills test requiring minimal time and resources is needed. Therefore, we set out to develop and validate a test for routine use in busy, low-resource maternity units; to efficiently and effectively assess potential knowledge and skills gains over time when attending ‘low-dose, high-frequency’ training in integrated care during childbirth.

**Materials and Methods:** Using Messick’s standards, we developed a comprehensive yet time-efficient test covering childbirth surveillance, respectful care, and management of maternity and neonatal complications. Both expert and participant feedback informed the test design. The test was applied during a Tanzanian in-house training intervention (PartoMa) to assess its performance. Reliability was determined by Cronbach’s Alpha, while validity was evaluated through face and content validity, factor analysis, and Rasch analysis.

**Results:** After multiple revisions, Cronbach’s Alpha was below 0.7, indicating limited reliability. Experts agreed the test was well-designed for the intended content, with no concerns about clarity or relevance. Content validity was confirmed through expert judgment, reflecting the test’s goals. In pilot testing, 160 (84.6%) participants rated it excellent in achieving the study objectives. Exploratory factor analysis showed the test did not measure a single latent trait, and Rasch analysis revealed discrepancies between observed and expected responses on item scores.

**Conclusion:** The knowledge and skills test has shown promise for assessing healthcare providers in high-pressure, resource-limited settings, but its low Cronbach’s Alpha and limitations in test results highlight the need for refinement.

## Introduction

An estimated 300,000 women lose their lives each year during pregnancy and childbirth globally. In addition, 2.7 million stillbirths and neonatal deaths occur and many more suffer from morbidity and lifelong disabilities (1). This burden is particularly severe in sub-Saharan Africa and Southern Asia where intra-facility quality of care has not followed suit with increasing numbers of facility births (2). Frontline healthcare providers are the essential agents for improving intra-facility quality of care. In resource-constrained settings, there is, however, often doubt about the capabilities of skilled birth attendants to provide quality care. Insufficient expertise among the limited number of birth attendants is a key reason for the need for ongoing training (3). Traditionally, such training has been carried out as lengthy, one-time training sessions (4). This method involves intensive training sessions to equip healthcare professionals with skills for emergency obstetric and newborn care.

In contrast to other approach, recurring in-house training workshops are increasingly being recommended as an effective approach for enhancing competencies in both low- and high-resource clinical settings (5,6). The ‘low-dose, high-frequency’ training model is gaining attraction as a strategy to enhance the retention of clinical knowledge, skills, and attitudes. This approach involves recurring, brief, in-house sessions focused on competencies, using simulation and case-based learning activities that are spaced out and built upon over time. The training emphasizes essential information and is often multidisciplinary to promote effective teamwork (7). This approach is backed by evidence indicating that: 1) recurring, targeted training is linked to better retention of knowledge and skills; 2) conducting training in the same environment as clinical practice leads to greater skills improvement, better performance and reduced costs compared to off-site, one-time training; 3) interactive methods such as simulation and case-based learning are more effective than traditional lectures; and 4) multidisciplinary team training increases the likelihood of translating knowledge and skills into clinical practice (3,8,9). The use of validated and suitable tools for measuring the knowledge and skills of frontline health workers is essential to ensure safe maternal care during labor and birth (10). Such tools not only validate the effectiveness of training programs but also contribute to consistent, evidence-based improvements in maternal health outcomes (11). Without these tools, it becomes difficult to quantify progress, allocate resources appropriately, or make necessary policy adjustments to support frontline health workers in their critical roles. Hence, there is a crucial need for suitable and validated assessment tools that align with the specific contextual requirements to measure potential improvements in knowledge and skills (12,13).

We aimed to develop a valid, low-resource knowledge and skills test that reflects real-world childbirth care in busy Tanzanian maternity units. We applied for the ongoing low-dose, high-frequency training of the PartoMa project. The Zanzibar PartoMa project is a capacity-building initiative aimed at improving maternal and neonatal health outcomes by providing context-specific intrapartum care guidelines and participatory training seminars for healthcare providers in resource-limited settings(6). It has demonstrated success in enhancing clinical decision-making, increasing the use of partographs, and improving maternal and neonatal survival rates through cost-effective, locally tailored interventions(14).

In a pilot intervention study conducted in Zanzibar, the PartoMa team observed that utilization of in-house, low-dose, high-frequency training in combination with co-created clinical practice guidelines (the PartoMa intervention) appeared to improve knowledge and skills and was associated with enhanced childbirth care and survival rates (6,15).

The knowledge and skills test used to evaluate the birth attendants who attended the PartoMa training was not validated and was thought to be insufficient. This study aimed to describe the participatory process of developing and validating a suitable and comprehensive knowledge and skills test to assess the impact of knowledge changes on the birth attendants who attended the seminars. As such, this validation process might serve as an exemplary to others developing similar knowledge and skills tests for clinical interventions in resource-limited settings.

## Methodology

### Setting

This development and validation study was conducted in Zanzibar, a semi-autonomous region of Tanzania with a population of 1,889,773 according to the final 2022 census report(16). Data were gathered during ongoing PartoMa seminars, primarily held in Mnazi Mmoja Hospital, the main tertiary hospital in Zanzibar. Seminar participants were drawn from various hospitals across Zanzibar that provide maternity care. Zanzibar’s Mnazi Mmoja Hospital facilitates approximately 11,500 births annually and accounts for 33% of all facility births in Zanzibar (17). With 30 to 50 daily births, it operates with a low staff level, relying heavily on junior doctors for intrapartum monitoring due to a nurse/midwife to laboring women ratio of 1:4 (18),(19). Mnazi Mmoja Hospital serves as a referral and training centre, playing a major role in teaching future healthcare professionals (20,21).

In Zanzibar, skilled birth attendants provide maternal care during pregnancy, labor and postnatal period (22). These health professionals are generally midwives, nurses or doctors with specific skills in managing uncomplicated pregnancies and births. They are also skilled in recognizing and responding to complications that may arise during labor, ensuring both the safety of the woman and her baby (23–25). Professional experience spans a spectrum from recent graduates to experienced practitioners with varied backgrounds. Educational qualifications range from diploma-level certifications to master degrees in relevant health disciplines (26). Continuous professional development is supported through refresher courses and on-the-job training to keep evidence-based practices (27). Challenges, however, persist due to limited access to up-to-date medical resources, inadequate infrastructure and shortage of qualified personnel hindering adaptation in resource-constrained environments to evolve healthcare practices (28). While the official medical language in Zanzibar is English, Swahili is mainly spoken in the health sector.

### Study design

The development of the knowledge and skills test followed an iterative process, guided by validation standards set by Samuel Messick and the practitioner’s guide of Goldstein and Behuniak,(29). Messick’s frameworks describe construct validity with several aspects, including content, substance, structure, external validity, and impact of a test. Goldstein and Behuniak provided a comprehensive framework for practitioners working within specific professional fields. This guide serves as a practical tool designed to bridge theoretical knowledge with real-world applications, helping professionals implement effective strategies and interventions. The development and validation processes of the PartoMa knowledge and skills test followed these standards across two phases (please refer to Figure. 1 of supplementary files). Phase One consisted of an iterative process of the development, validation, and reliability assessment of the test. Phase Two included revisions, pilot testing, and evaluations of the test.

**Figure 1.**
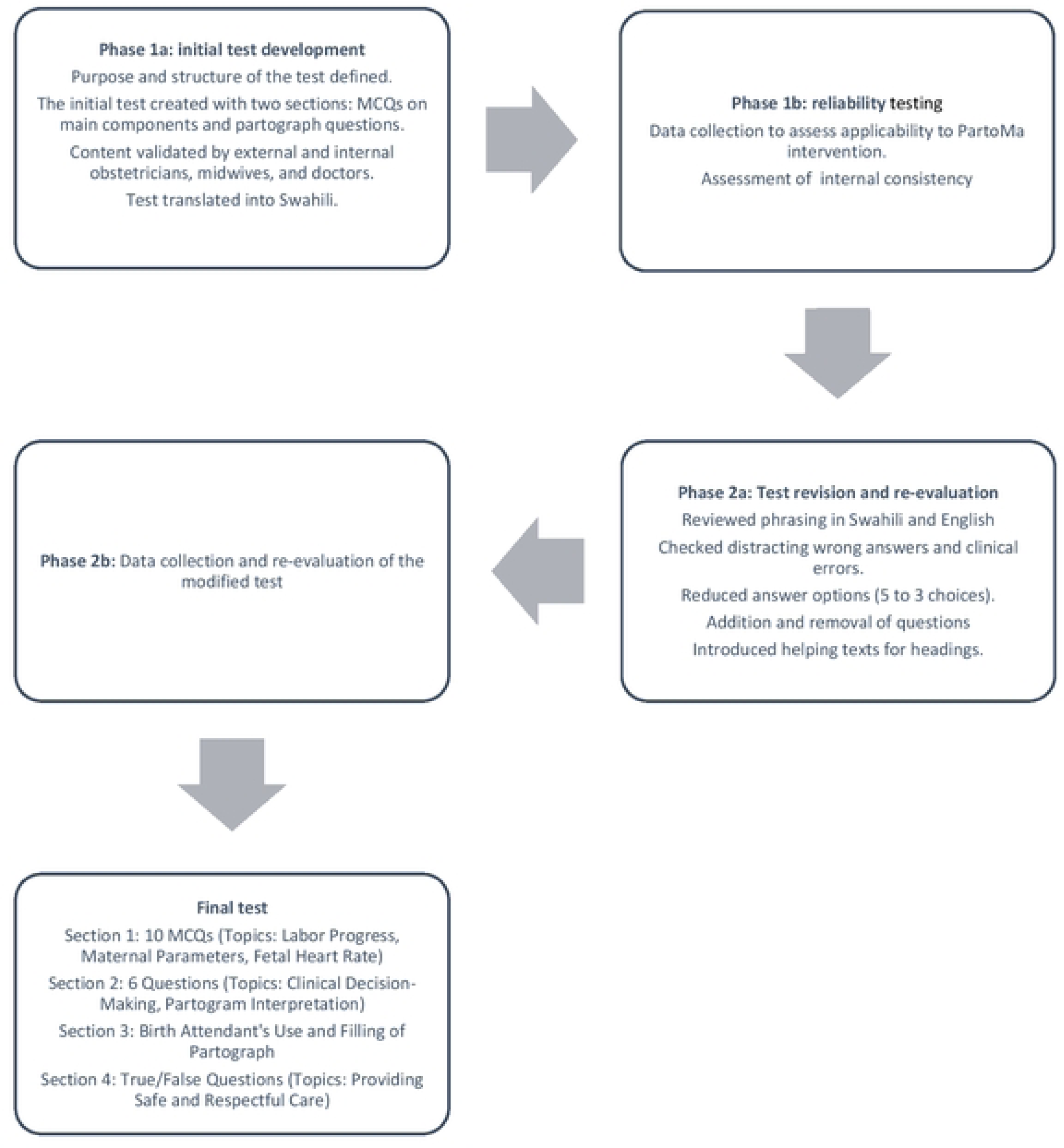
PartoMa knowledge-skil Test Development and validation Process.

### Phase One: Initial development, content and face validation, and reliability

The initial PartoMa knowledge and skills test was developed during the PartoMa pilot study in Zanzibar in 2016 by NM with input from specialists in obstetrics, midwives and senior researchers(15). This was followed by a validation process guided by MH, an expert in research tool validation. The test focused on essential clinical knowledge aligned with WHO’s intrapartum care domains(30). Utilizing the “one-best-answer principle”, the initial test had two sections, including multiple-choice questions (Section One) and partograph completion and interpretation tasks (Section Two). The test was in both English and Swahili, as well as designed for self-administration (please refer to Annex 1 in the supplementary files). To ensure its validity, two external obstetricians, one external midwife and two internal doctors from Mnazi Mmoja Hospital, all knowledgeable about the PartoMa intervention, reviewed the test for content and face validity. Modifications were made based on their feedback. They evaluated the test’s applicability for assessing learning essential clinical knowledge for integrated safe and respectful care during childbirth. The reviewers (all with clinical experience in obstetrics) clarified the test’s phrasing in Swahili and English and determined whether the distracting wrong answers fell into the same category as typical clinical errors. They were encouraged to focus on the content’s accuracy and consider the clinical relevance of the test, as well as its generalizability to other settings.

### Phase Two: Pilot testing and further refinement

During Phase Two in 2022, the PartoMa team collaborated with local senior doctors and midwives to conduct pilot testing of the revised modified test after incorporating the suggested changes from the initial analysis. Helping texts were introduced for headings, and the number of answer options was reduced from five to three. Data from the pilot testing was obtained from PartoMa seminar participants, who were skilled birth attendants in different hospitals across Zanzibar providing maternity care (please refer Anex 2). In this testing, an evaluation questionnaire also gathered participants’ opinions on relevance, content, and usability, graded on a scale of poor to excellent (please refer to Annex 3).

### Data collection

Data collection was done during the seminars conducted in-house as ‘low-dose, high-frequency training’ offered once every three months during both 2016 and 2022. Following the pilot intervention there was a stop of active PartoMa seminars that halted the use of the test and disrupted its further development and validation process. For the revised test, data were collected during the implementation period from 2022 to 2023. Only the pre-test (pre-seminar) data were included in the present study because our goal was to measure the validity of the knowledge and skills test. Participants who had previously attended PartoMa seminar were excluded from this study. According to the literature, sample determinations can be based on the respondent-to-item ratio(31). It is recommended to have between 5 to 15 participants for each item. Therefore, the PartoMa test, which consisted of 22 items, required a total of 110 to 330 respondents. To minimize bias in participant scores, we advised participants to avoid skipping questions and we removed the ‘no answer’ option, as that could significantly reduce the overall numbers of responses(31).

### Statistical analysis

Statistical analyses were performed using the statistical software packages SPSS (version 28.0) and R(32). First, simple descriptive analyses using frequency distributions were used to describe the demographic characteristics of study participants regarding age, workplace, employment professional, and work experience. Test scores were presented as a means with standard deviations and stratified according to work experience and employment groups for the two different sections of the test. Internal consistency of the initial test was analyzed using CA on a scale of 0 to 1.0. A high value of the CA score indicates that the different items within the test measure the same concept. CA scores >0.7 indicate adequate internal consistency and scores ≥ 0.9 suggest excellent internal consistency (12). To ensure comprehensive assessment, it was crucial to measure outcomes not only for the entire test but also for the individual sections, as each section measured different concepts. Thus, CA scores were calculated as follows: 1) for the complete test, with each question worth 1 point; 2) for the complete test, which included the weighted scores for section two and 3) for each section of the test. CA was also calculated for subgroups of participants stratified by profession and experience as these could potentially influence internal consistency. Lastly, a set of validity and reliability calculations was done to assess whether removal of specific questions would affect internal consistency. A one-way ANOVA was used to determine whether test scores between the subgroups were different, using a p-value <0.05 for statistical significance.

After revision, validity and reliability checks were done using exploratory factor analysis and Rasch analysis (32,33). Exploratory factor analysis was used to assess the number of dimensions the test covers (34). This was done by visual examination of screen plots (Please refer to Annex 4 in supplementary material). Rasch analysis was performed using visual comparison of observed and expected mean scores across different groups’ total scores (supplementary Annex 5-8). The marginal distribution of item scores was visualized using the R package RASCH plot(35).

### Ethical considerations

Permission to conduct this study was obtained from the Zanzibar Health Research Institute (ZAHRI) with permit numbers (ZAMREC/0001/JUNE/014, 7th October 2014) for the pilot version, and current version permit number (ZAHREC/04/ST/JULY/2021/60). The participants involved in the knowledge and skills test were informed about the study and gave consent which they were free to withdraw at any time. Additionally, they were also assured of confidentiality in the management and storing of data, as well as in the reporting of the results.

## Results

### Development of the knowledge and skills test

When the test was first analyzed, Cronbach’s Alpha (CA) was found to be low (<0.7), prompting suggested changes to improve reliability. The changes included removing certain questions, modifying existing questions, and adding new questions to enhance the research tool’s reliability. The final version of the knowledge and skills test included 22 questions with 27 test items in total broadly covering safe and respectful routine and emergency care during childbirth (Annex 2). There are four sections in the test. The first section included ten multiple-choice questions, each with three answering options and one correct answer.

Additionally, this section focused on aspects related to fetal heart rate, maternal parameters, and labor progress. In the second section, which consisted of six questions, partograph interpretation, and clinical decision-making were addressed as skills-related components. The third section focused on accurate plotting of data on the partograph. The fourth section addressed the provision of safe and respectful care for expectant mothers with a true/false response option. The clinician-researchers in the team unanimously agreed that the proposed solutions were the optimal responses.

### Background characteristics of the seminar participants included in phase one

Table 1 provides an overview of the study population’s demographic characteristics. A total of 136 birth attendants participated in the study. More than two-thirds of 97 (71.3%) were in the age group 20-29 years. About half of them 67 (49.5%) were employed in Mnazi Mmoja Hospital, and more than half 80 (58.8%) had work experience of less than a year. The group of nurse-midwives and medical officers, who have a diploma level of education in Tanzania referred to as clinical officers, comprised the largest proportion of all study participants 58 (42.6%). The group of medical and nursing students reported having little work experience, as nearly the entire group had less than a year of work experience. Therefore, students’ test scores were not divided into subgroups based on work experience. Most participants, 80 (58.8%), had less than one year of work experience.

**Table 1.**
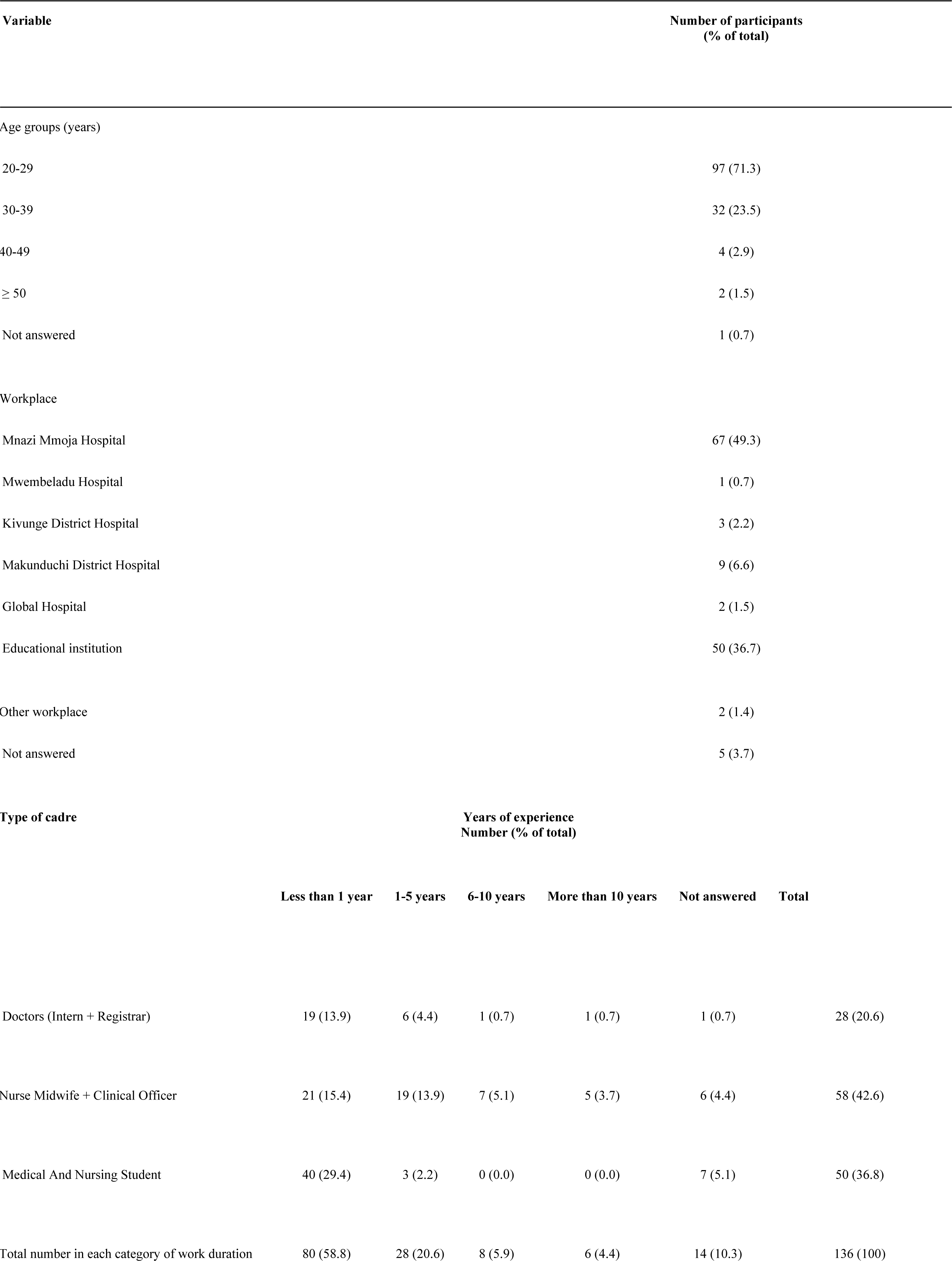
Demographic characteristics of participants (N = 136).

Table 2 provides an overview of mean test scores according to profession and duration of work experience. The total mean score among participants was 10.36(SD 2.50). Medical doctors presented with the highest means 11.22 points (S.D 2.47), and nurses’ midwives with the lowest means 9.12 points (SD 2.44). The highest test score was achieved by a medical doctor with 6-10 years of work experience (16.00 points), whereas the lowest was achieved by a nurse with 6-10 years of work experience (7.75 points). There was only a small difference between scores across occupational status and duration of work experiences.

**Table 2.** Knowledge and skills test score results among participants according to profession and duration of work experience in years (N=136). The initial test comprised two sections. The table presents the scores for both sections.

| Occupation (n) | Section One mean scores<sup>†</sup> | Total test mean scores<sup>††</sup> |
| --- | --- | --- |
| Doctors (Intern + registrar (n=28)) | 9.21 (S.D. 2.06) | 11.22 (S.D. 2.47) |
| Nurse midwife + clinical officer (n=58) | 7.26 (S.D. 2.05) | 9.12 (S.D. 2.44) |
| Medical and nursing student; (n=50) | 8.72 (S.D. 1.72) | 11.00 (S.D. 2.15) |
| Duration of work experience for all participants;<br>in years (n) |  |  |
| <1 (n=80) | 8.58 (S.D. 1.95) | 10.93 (S.D. 2.19) |
| 1-5 (n=28) | 8.00 (S.D. 1.85) | 9.88 (S.D. 2.22) |
| 6-10 (n=8) | 6.88 (S.D. 3.00) | 9.40 (S.D. 4.16) |
| >10 (n=10) | 9.00 (S.D. 1.90) | 12.00 (S.D. 1.41) |
| Duration of work experience for medical doctors;<br>in years (n) |  |  |
| <1 (n=19) | 8.74 (S.D. 1.94) | 10.78 (S.D. 2.56) |
| 1-5 (n=6) | 10.00 (S.D. 1.55) | 11.83 (S.D. 1.33) |
| 6-10 (n=1) | 13 | 16 |
| >10 (n=1) | 12 | 13 |
| Duration of work experience for nurses; in years (n) |  |  |
| <1 (n=21) | 7.76 (S.D. 2.45) | 9.94 (S.D. 2.52) |
| 1-5 (n=19) | 7.37 (S.D. 1.61) | 9.18 (S.D. 2.27) |
| 6-10 (n=7) | 6.00 (S.D. 1.83) | 7.75 (S.D. 2.22) |
| >10 (n=5) | 8.40 (S.D. 1.34) | 11.00 (S. D=1.32) |
| All participants (n=136) | 8.20 (S.D. 2.09) | 10.36 (S.D. 2.50) |

<sup>i</sup> Highest possible score that a test-taker can achieve in Section One (in this section a maximum of 13 points).
<sup>ii</sup> Total test score refers to the average score achieved by all test-takers who have completed a test (In this test, the maximum score is 16 points).

### Validation (Cronbach’s Alpha, exploratory factor analysis and Rasch analysis)

An overview of the proportion of respondents who indicated a correct answer for each question is presented in Table 3. Questions 6 and 9 were about maternal sepsis in labor and answered poorly by all groups as was question 9, which was about the skills to read and interpret a partograph that showed fetal distress. Question 2 about non-re-assuring fetal heart rate was answered very well by all groups. Question 6 had a result opposite of what would be expected from table 3, where nurses midwives answered this question better than the group of doctors.

**Table 3.**
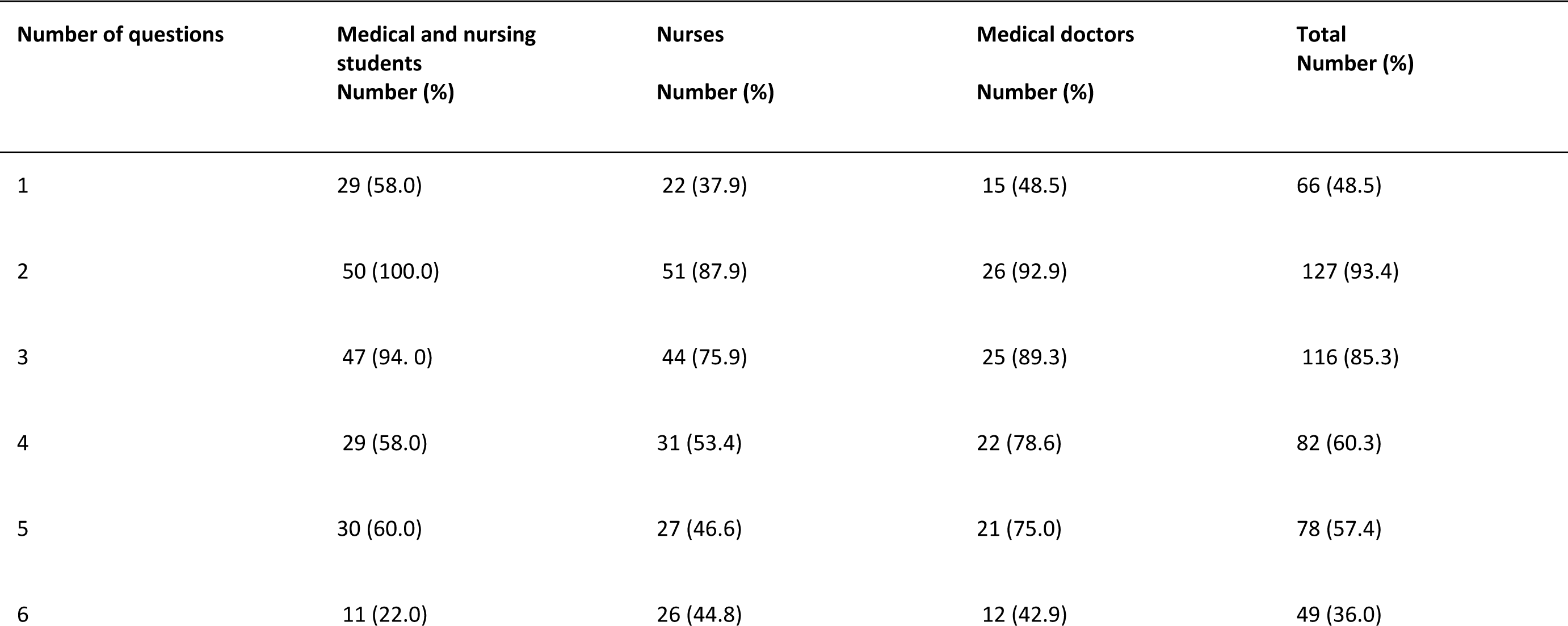
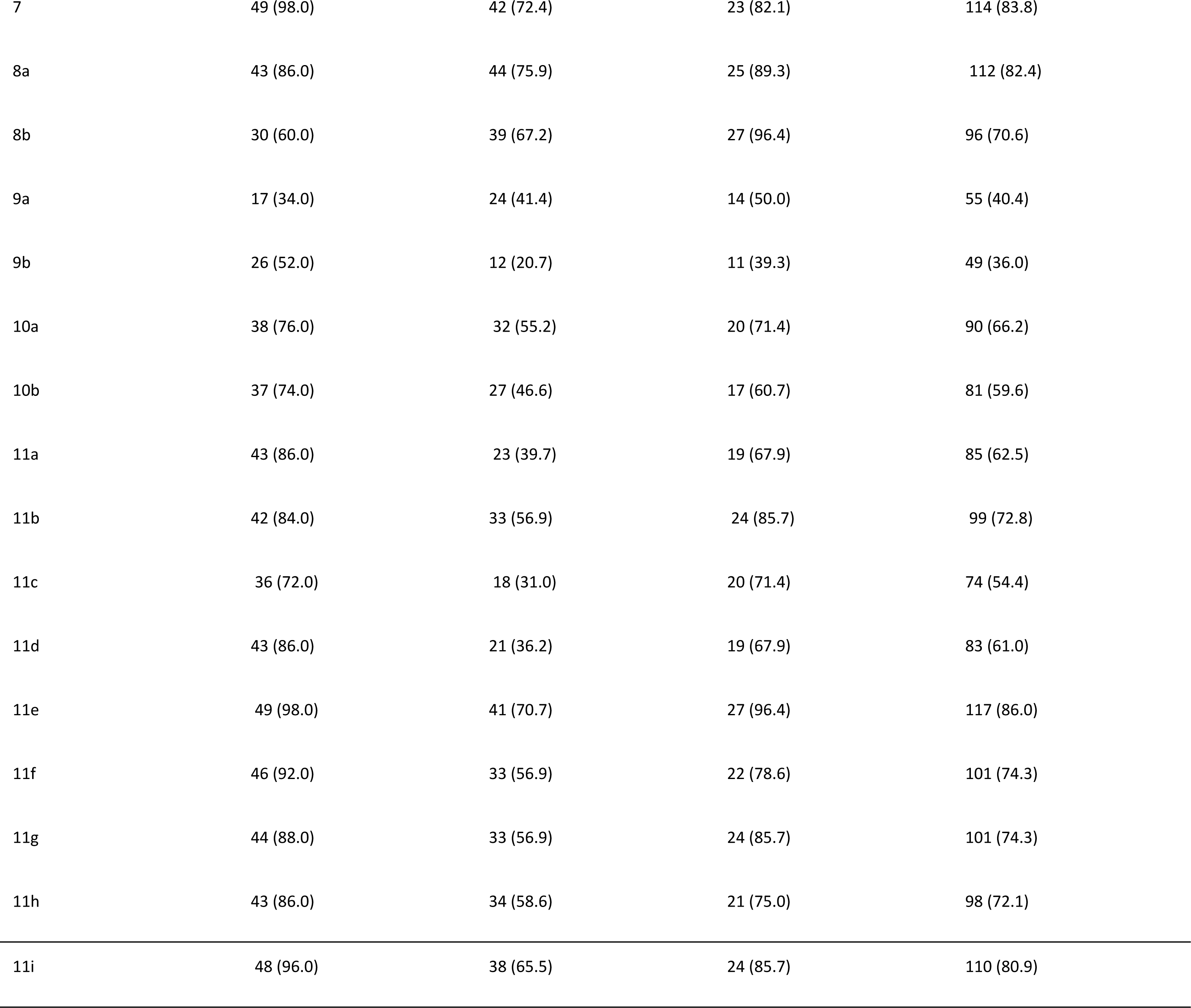
Proportion of participants who answered each question correctly (N=136).

The results of the analysis of using CA for the initial test are shown in Table 4. CA for reliability was 0.54 in the whole group, below the required 0.70. The lowest CA score was 0.25 (Section One for nursing students with 1-5 years of work experience), whereas the highest score was 0.74 (Section Two; for the total participants).

**Table 4.**
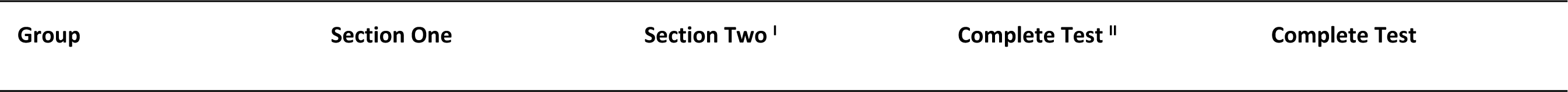
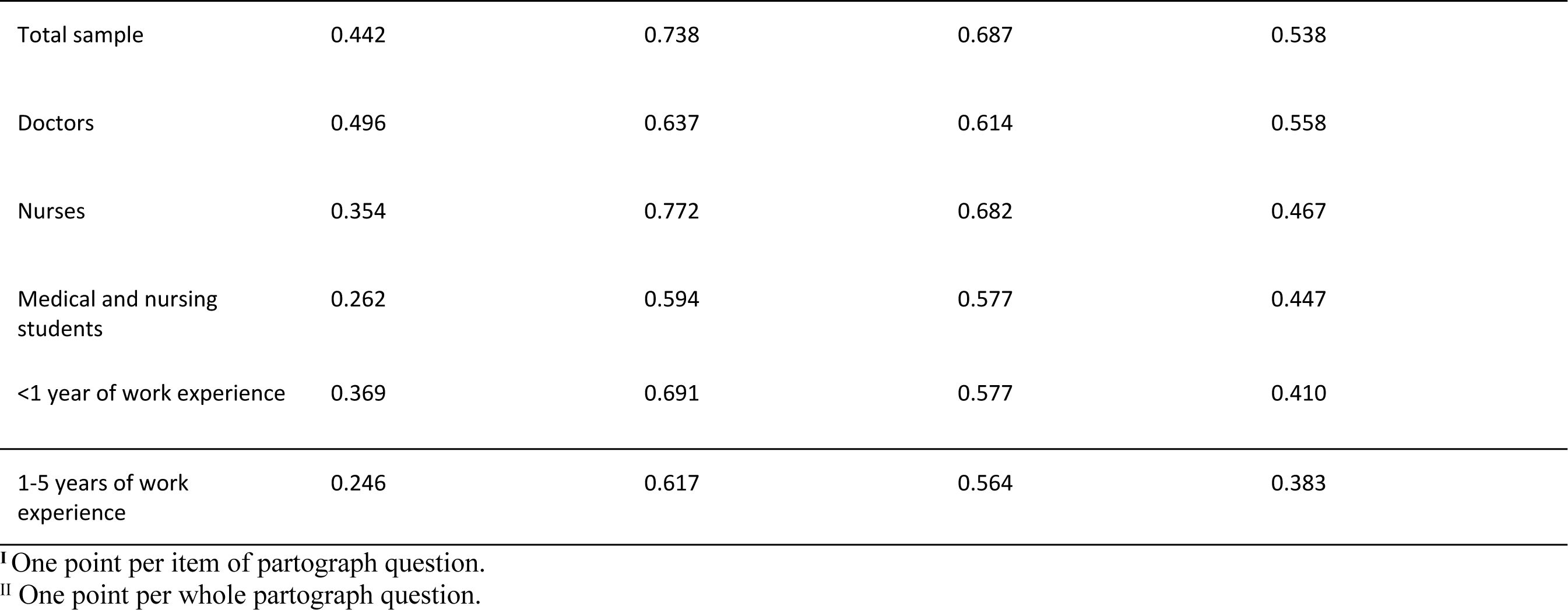
Cronbach’s Alpha values (N=136). The initial test had two sections: Section One multiple-choice questions; Section Two assessment of the use of partograph.

| Group | Section One | Section Two <sup>i</sup> | Complete Test <sup>ii</sup> | Complete Test |
| --- | --- | --- | --- | --- |
| Total sample | 0.442 | 0.738 | 0.687 | 0.538 |
| Doctors | 0.496 | 0.637 | 0.614 | 0.558 |
| Nurses | 0.354 | 0.772 | 0.682 | 0.467 |
| Medical and nursing students | 0.262 | 0.594 | 0.577 | 0.447 |
| <1 year of work experience | 0.369 | 0.691 | 0.577 | 0.410 |
| 1-5 years of work experience | 0.246 | 0.617 | 0.564 | 0.383 |

<sup>I</sup> One point per item of partograph question.
<sup>II</sup> One point per whole partograph question.

### Phase two results

#### Demographic characteristics of the participants

The demographic characteristics presented in Table 5 are respondents of pilot testing. This table presents the results from rounds 1 to 3 of the pilot testing of the PartoMa test.

**Table 5.** Demographic characteristics of participants who performed pilot testing (N=189).

| Employment | Number of participants in pilot seminars (% of total) |  |  |
| --- | --- | --- | --- |
|  | Interview round 1 (n=31) | Interview round 2 (n=38) | Interview round 3 (n=120) |
| Intern doctor |  |  |  |
| Doctor | 0 (0) | 1 (2.6) | 20 (16.7) |
| Nurse | 7 (22.6) | 3 (7.9) | 16 (13.3) |
| Clinical Officer | 21 (68.1) | 29 (76.3) | 82 (68.4) |
| Assistance Medical Officer | 2 (6.4) | 2 (5.3) | 0 (0) |
| Other Employment | 1 (3.2) | 2 (5.3) | 1 (0.8) |
| Total | 0 (0) | 1 (2.6) | 1 (0.8) |
|  | 31(100) | 38(100) | 120(100) |
| Workplace |  |  |  |
| Mnazi Mmoja Hospital | 0 (0) | 0 (0) | 43 (35.8) |
| Kivunge District Hospital | 21 (68.1) | 34 (89.5) | 0 (0) |
| Makunduchi District Hospital | 2(6.4) | 0 (0) | 0 (0) |
| Other facility from North region | 8 (25.8) | 4 (10.5) | 8 (6.7) |
| Other facility South region | 0 (0) | 0 (0) | 11 (9.2) |
| Abdallah Mzee Hospital (Pemba) | 0 (0) | 0 (0) | 0 (0) |
| Another workplace | 0 (0) | 0 (0) | 58 (48.3) |
| Not answered | 0 (0) | 0 (0) | 0 (0) |
| Experience with maternity care (years) |  |  |  |
| <1 | 11 (35.5) | 2 (5.3) | 39 (32.5) |
| 1-5 | 10 (32.3) | 24 (63.2) | 76 (63.3) |
| 6-10 | 5 (16.0) | 9 (23.7) | 4 (3.3) |
| >10 | 5 (16.0) | 3 (7.9) | 1 (0.8) |
| Age (years) |  |  |  |
| 20 - 29 | 17 (54.8) | 18 (47.4) | 2 (1.7) |
| 30 – 39 | 8 (25.8) | 16 (42.1) | 111 (92.5) |
| 40 – 49 | 4 (12.9) | 1 (2.6) | 6 (5.0) |
| 50 and above | 2 (6.5) | 3 (7.9) | 1 (0.8) |

Participants included midwife nurses, doctors, intern doctors, clinical officers, assistant medical officers, and various other healthcare professionals. Throughout all three rounds of testing, nurses represented the largest group among participants, accounting for 21 (68.1%), 29 (79.3%) and 82 (68.4%). Most participants were employed in Kivunge and other healthcare facilities in the northern region, with figures of 21 (68.1%) and 34 (89.5%) in two of the three rounds.

In terms of experience in maternity care, most participants had less than one year of experience across all three rounds, with totals of 10 (32.3%), 24 (63.2%), and 76 (63.3%). Additionally, most participants were within the 30-59 years age group 111 (92.5%).

### Participants’ perceptions of the test and test response time

Table 6 provides an overview of the views and opinions of the participants regarding questionnaire design and whether it covers the relevant aspects of intrapartum care. The majority rated the test design as “excellent” or “good”. The fastest participant completed the test in 20 minutes, while the slowest took 28 minutes.

**Table 6.** Participants’ opinions about test design and content (N=189). Column one displays available options for the test category. Other columns show respondents’ numbers and percentages (in brackets) for each test category.

| Participant's grading of each dimension of a questionnaire | The average style of the questionnaire<br>Number (%) of total | Coverage of the knowledge of the field<br>Number (%) of total | Relevancy and necessary<br>Number (%) of total | Length for reducing Burden to the respondent to fit with the time planned<br>Number (%) of total | Flows seamlessly from one question to the next to follow a logical and intuitive layout that for the same sake reduces respondent burden<br>Number (%) of total | The first-contact response rate (whether the response rate after the first contact meets expectations) as well as response rates for subsequent contacts<br>Number (%) of total | The first debriefing of the researcher about the test, how do you value it?<br>Number (%) of total | Your final conclusion about the test in relation to the intervention of the researcher, how do you grade it?<br>Number (%) of total |
| --- | --- | --- | --- | --- | --- | --- | --- | --- |
| Excellent | 90 (47.6) | 120 (63.4) | 88 (46.5) | 99 (52.3) | 134 (70.8) | 150 (79.3) | 179 (94.7) | 160 (84.6) |
| Good | 40 (21.2) | 40 (21.1) | 65 (34.3) | 70 (37.0) | 45 (23.8) | 20 (10.5) | 8 (4.2) | 24 (12.6) |
| Average | 52 (27.5) | 19 (10.0) | 25 (13.2) | 11 (5.8) | 8 (4.2) | 17 (8.9) | 2 (1.0) | 5 (2.6) |
| Poor | 7 (3.7) | 10 (5.2) | 11 (5.8) | 9 (4.0) | 2 (1.0) | 2 (1.0) | 0 (0.0) | 0 (0.0) |
| Total | 189 (100) | 189 (100) | 189 (100) | 189 (100) | 189 (100) | 189 (100) | 189 (100) | 189 (100) |

Rasch analysis was performed on test scores from 189 participants. This analysis, detailed in Annex 5 for visualization of items, and Annex 6-8 for Rasch analysis models, allowed us to observe how the questionnaire items were answered and to compare these responses with the expected answers based on the Rasch measurement model. It made it possible to assess the extent to which the questionnaire items measured the intended construction, pinpoint problematic items, and evaluate the overall quality of the questionnaire. Exploratory factor analysis indicated multi-dimensionality (Annex 4) and the Rasch analysis revealed significant differences between observed and expected item response scores (Annex 6-8).

## Discussion

This study aimed to develop and validate a concise, practical knowledge and skills test for assessing safe and respectful routine and emergency maternity care during childbirth. The finalized instrument consists of 22 multiple-choice and true/false questions, targeting critical competencies such as fetal monitoring, partograph interpretation, and clinical decision-making. Designed to be completed within 30 minutes, the tool was tailored specifically to align with the content of the PartoMa guidelines and training initiative in Tanzania. Tools of this nature are essential for identifying and addressing barriers to providing appropriate and respectful maternity care (36). As part of a broader intervention adapting WHO guidelines for busy maternity units, the test is intended to comprehensively address intrapartum care(12,37,38).

It has been emphasized that a tool’s validity is maximized when it comprehensively addresses the intended competencies and aligns with the goals of the applied intervention(39–41).

Despite the tool’s alignment with study objectives and its intended relevance, statistical validation tests revealed that the PartoMa knowledge and skills test did not measure a unidimensional latent trait. This finding indicates challenges in capturing cohesive maternity care knowledge and skills construct. These results highlight the inherent difficulties in creating statistically validated tests that remain practical and feasible in real-world settings(42). Nonetheless, feedback from end-users and experts confirmed the tool’s perceived relevance and utility.

This study contributes to the existing body of knowledge by developing a tool specifically aligned with the PartoMa training, bridging the gap between theoretical guidelines and practical implementation(34,43–47). Several studies highlight the importance of developing and validating assessment tools to evaluate knowledge and practices across various healthcare domains(48–55). Key insights from these cited studies include three main areas:

(1) enhancing the skills of frontline healthcare providers to improve patient outcomes; (2) ensuring methodological rigor through the use of validated, reliable, and culturally relevant tools, and (3) diverse topics such as evidence-based practice, palliative care, and health literacy. The present PartoMa validation study aligns with this approach by providing a comprehensive evaluation of maternity care competencies among frontline healthcare providers.

Unlike many other tools, which often lack context-specific customization, the PartoMa study knowledge-skills test was explicitly designed for Tanzanian maternity units, addressing the unique challenges of resource-limited and high-pressure environments(56). There are several distinct differences when comparing the PartoMa validation study with other similar studies(48,53–55). Firstly, the present study offers a dual assessment strategy, integrating both routine and emergency maternity care within a single evaluative framework. This approach effectively addresses a notable gap in existing assessments, which predominantly focus on routine practices(51,57–59). Secondly, the present study places a strong focus on critical care. Prioritizing emergency maternity care underscores the importance of swift and accurate decision-making during childbirth, a critical factor in reducing maternal and neonatal mortality. Thirdly, the PartoMa validation study is designed for direct application in resource-limited settings, enhancing its relevance for regions plagued by high maternal and neonatal mortality rates. Lastly, it promotes collaboration among frontline cadres by assessing a diverse group of healthcare providers, including doctors, nurses, and midwives, in a collective effort to advance team-based maternity care.

### Strengths and limitations

The study’s strengths lie in its comprehensive validation framework, utilizing Messick’s approach to ensure alignment with widely accepted standards. The test was specifically tailored to the competencies addressed in the PartoMa training intervention, enhancing its relevance and applicability. Expert reviews affirmed content validity, while pilot testing showed that 84.6% of participants felt the test met its objectives, highlighting its utility among frontline healthcare workers.

Despite its strengths, the study encountered several limitations. Reliability analysis indicated a low Cronbach’s alpha, suggesting the need for refinement to improve internal consistency. Factor and Rasch analyses revealed concerns with one-dimensionality, indicating that the test may not fully capture a single underlying construct. Specific test items deviated from expected response patterns, requiring item-level refinements to improve clarity and relevance. Additionally, the test’s performance and generalizability need to be evaluated in other settings.

Future studies should focus on refining the test items to enhance reliability and clarity. Revisiting the number of test items and conducting item-level analysis could improve internal consistency. Additionally, testing the tool across diverse settings will help assess its generalizability and broader applicability. Future research should also explore digital adaptations of the test for scalability and usability in high-pressure environments.

This study contributes to maternity care by offering a validated, context-specific tool for assessing intrapartum care competencies. It addresses a critical gap by integrating routine and emergency obstetric care into a single evaluation framework. The test’s emphasis on brevity and applicability within resource-limited settings underscores its potential to improve maternal and neonatal outcomes. By promoting collaboration among healthcare cadres, the tool advances team-based maternity care, aligning with global efforts to reduce maternal and neonatal mortality(60,61).These perspectives underscore the importance of promoting team-based maternity care to enhance maternal and neonatal health outcomes.

### Conclusion

This study successfully developed a knowledge and skills test aligned with the objectives of the PartoMa training intervention, confirming its content validity through expert and user feedback. However, challenges in internal consistency and dimensionality highlight the need for further refinement. Addressing these limitations will improve the tool’s reliability and its capacity to accurately measure frontline workers’ competencies in delivering safe and respectful maternity care. Despite these challenges, the tool represents an innovative advancement in enhancing maternity care outcomes in resource-limited settings.

## Data Availability

The data supporting this study are available in our institution repository. Currently, the data are stored in our institutional repositories, which are accessible only to the authors. Our institution employs a specialized system for securely managing and storing both sensitive and non-sensitive data, in compliance with the General Data Protection Regulation (GDPR). Access to these data can be granted, subject to institutional guidelines and ethical considerations.

## Acknowledgments

The research team extends thanks to healthcare professionals in the relevant facilities who actively participated in responding to the test. This includes Mnazi Mmoja Hospital, peripheral district hospitals of North and South Unguja, and maternal delivery health facilities in Pemba Island. We also acknowledge the cooperation of hospital and maternity department heads, who allowed their staff to participate and provided venues for the test. Special appreciation is reserved for Suhaila Salum Yusuf, the research assistant, for her dedicated efforts in ensuring the timely administration of the test at all pilot testing locations and for meticulously collecting and organizing questionnaire responses for subsequent analysis.

